# Real world data analysis of venous thromboembolism, adverse major cardiovascular events, neoplasia and serious infections in ulcerative colitis patients

**DOI:** 10.1101/2025.01.21.25320903

**Authors:** Víctor de la Oliva, Alberto Esteban-Medina, Patricia Fernández del Valle, Ana Sánchez, M. Belen Susin, Joaquín Dopazo, Carlos Loucera, Eduardo Leo-Carnerero

## Abstract

Venous thromboembolism (VTE), major adverse cardiovascular events (MACE), neoplasia, and serious infections are significant complications associated with Immune-mediated inflammatory diseases. Actually, the occurrence of such complications in these patients has been observed to be higher than that anticipated from classical risk factors alone. Additionally, the administration of anti-inflammatory treatments, such as corticosteroids, associated to inflammatory diseases is recognized to further elevate this risk. In Spain, data on the prevalence of risk of cardiovascular events, neoplasia or serious infections, particularly among individuals with ulcerative colitis are scarce. This study leverages real-world data from the Andalusian Health Population Database (BPS) to analyze the incidence and risk factors of these complications among ulcerative colitis patients. A cohort of 23,518 patients, aged 18 years or older, with an ulcerative colitis diagnosis between 2010 and 2019, was used in the study. The objective was to assess the incidence of VTE, MACE, neoplasms, and serious infections and to evaluate the impact of age and other factors related to ulcerative colitis on these outcomes.

The study revealed a notably higher incidence of VTE and MACE in ulcerative colitis patients compared to the incidence described for the general population, particularly among those over 60 years of age. Specifically, the incidence of VTE was significantly elevated post-diagnosis, with deep vein thrombosis (DVT) and pulmonary embolism, being the most common manifestations. MACE, including myocardial infarction and stroke, also presented a higher risk, especially in older patients. Also, a significant portion of patients developed various malignancies. Also remarkable is the significant higher incidence of VTE, malignancies and serious infections as the severity of ulcerative colitis increases. The use of immunosuppressive therapies was associated with an increased risk of infections, and likely with VTE and malignancies, further complicating the management of these patients.

The findings of this study underscore the need for heightened vigilance in the management of ulcerative colitis patients, particularly those at advanced ages. Preventive strategies, such as thromboprophylaxis during hospitalization and regular monitoring for cardiovascular and neoplastic complications, are essential. The study contributes valuable insights into the burden of comorbidities in ulcerative colitis and highlights the importance of tailored treatment and monitoring strategies to improve patient outcomes.

## Introduction

Immune-mediated inflammatory diseases (IMIDs) can have a major impact on patients’ health, with cardiovascular risk, infections and the development of cancer being the long-term complications of these diseases that most concern both patients and physicians. Patients with IMIDs exhibit a cardiovascular risk that exceeds what would be anticipated based on traditional risk factors alone. Specifically, individuals with inflammatory bowel disease (IBD) have a heightened risk of both atherothrombotic and thromboembolic events, including coronary heart disease, arrhythmias, cerebrovascular accidents, and thromboembolism, compared to the general population. The risk profile varies according to the specific IMID: IBD is notably associated with an increased incidence of venous thromboembolism (VTE), whereas other inflammatory conditions are linked to a higher risk of early atherosclerosis and premature cardiovascular mortality. Additionally, the use of anti-inflammatory therapies, such as corticosteroids, has been implicated in elevating cardiovascular risk, while anti-TNF agents appear to mitigate this risk [1, 2].

Chronic inflammation plays a central role in increasing cardiovascular risk through mechanisms involving elevated cytokine levels, oxidative stress, platelet dysfunction, hypercoagulability, endothelial dysfunction [3], and, in the case of IBD, alterations in the gut microbiota [4]. Despite a lack of specific data on the prevalence of cardiovascular risk in Spanish patients with ulcerative colitis, numerous studies on IBD confirm an elevated cardiovascular risk, as highlighted in recent reviews [5]. Notably, this increased risk is particularly pronounced among women and young adults, resulting in higher incidence rates at younger ages [6] and contributing significantly to mortality among IBD patients [7].

Several studies indicate that patients with IBD may face an increased risk of malignancies, including leukemia, non-Hodgkin’s lymphoma, bile duct carcinoma, and bladder carcinoma [8, 9]. Immunosuppressive therapies used in conditions such as ulcerative colitis and atopic dermatitis may contribute to this elevated cancer risk. Evidence from studies involving solid organ transplant recipients suggests that immunosuppressants, particularly those used to prevent rejection, are associated with an increased risk of malignancy [10, 11], especially lymphoproliferative disorders [12, 13]. However, conflicting findings exist, as some studies in IBD patients have not demonstrated a clear link between immunosuppressant use and elevated cancer risk [14]. Emerging experimental and clinical data also suggest a potential causal relationship between infections and cancer development, although current evidence remains inconclusive due to limitations such as study duration and sample size [15]. Therefore, further research is essential to clarify these risks. Patients with IMIDs are also at higher risk for serious and opportunistic infections, particularly during immunosuppressive treatment and when multiple drugs (e.g., corticosteroids, immunomodulators, biologics) are combined. Improved strategies, including higher vaccination rates and enhanced infection screening, are needed to mitigate these risks in clinical practice [16–18].

Andalusia, the largest region in Spain and the third largest in Europe, boasts a population of 8.5 million, comparable to countries like Switzerland and Austria. The BPS, which contains medical records for over 15 million patients, represents a near-complete coverage of the Andalusian population [19]. This study leverages the vast amount of Real-World Data (RWD) available within the BPS, where detailed clinical information from 23,518 adults with an ulcerative colitis diagnosis between 2010 and 2019 were found. By utilizing this extensive dataset, the study aims to enhance awareness among physicians and patients regarding the risks associated with ulcerative colitis, allowing for timely intervention and improved management strategies. The findings will inform future clinical recommendations and contribute to the early detection and management of cardiovascular disease, neoplasms, and infections in ulcerative colitis patients, ultimately enhancing their quality of life. This research underscores the transformative potential of RWD [20] in refining healthcare practices and guiding decision-making in patient care, particularly for those with ulcerative colitis and related inflammatory conditions.

## Material and Methods

### Design of the study

This is a retrospective, multicenter, non-interventional, retrospective observational cohort study of patients diagnosed with ulcerative colitis using data included in the BPS.

The target population of the study were adult Andalusian patients with a diagnosis of ulcerative colitis (ICD10: K51) during the period 2010-2019, for whom data are available in BPS. Clinical data, along with sociodemographic variables, primary care, emergency and specialty care visits, hospitalization events, laboratory tests, comorbidities (heart disease, hyperlipidemia, history of malignancies, history of recurrent infections, extraintestinal manifestations, hypertension, cardiovascular history, diabetes), prescriptions, obesity, current smoker status and alcohol consumption, were requested.

The population of patients with moderate to severe ulcerative colitis is defined as those who in the study period were prescribed one of the following treatments: Corticosteroids (Beclomethasone, Budesonide, Deflazacort, Methylprednisolone, Prednisone, Prednisolone), Immunosuppressants (Azathioprine, Mercaptopurine, Methotrexate, Mycophenolate) or Biological treatment (Adalimumab, Golimumab, Ustekinumab, Infliximab, Vedolizumab) after their first ulcerative colitis diagnosis.

The following adverse events were studied: deep vein thrombosis (DVT), eliminating in both cases embolism and thrombosis of superficial veins; pulmonary embolism; venous thromboembolism (VTE), defined as patients who have had an episode of DVT and/or PE; major adverse cardiovascular events (MACE); malignant neoplasm except non-melanoma skin cancer; and severe infections. For all adverse events, except for severe infections, both outpatient and hospital admission diagnoses were considered. For severe infections, only hospital admission diagnoses were considered. All the ICD codes corresponding to the above-mentioned adverse events are listed in Supplementary Table 1.

### Sample size estimation

In conventional studies using hospital databases, the size calculation underlies the need to obtain a portion or subset of individuals with specific characteristics (i.e. sample) that is representative of the whole, and to be able to respond to the study objectives with adequate power and precision so that the results can be generalized to the whole population.

For the expected prevalence of 99.84 per 100,000 inhabitants [21], the required sample size would be 62,123 patients, for the margin of error or absolute precision of 0.02496% in estimating the prevalence with 95% confidence and considering the potential loss/attrition of 1%. With this sample size, the anticipated 95% CI is/was (0.07488%, 0.1248%). This sample size is calculated using the ScalaR SP [22]. This study uses Real World Data (RWD) and the major difference in this approach is that it encompasses the whole Andalusia population of 8.5 million inhabitants [23], including all patients who meet the selection criteria, i.e. the entire target population of ulcerative colitis patients in Andalusia.

### Statistical analysis

Incidences were calculated as the number of separate events, divided by the number of ulcerative colitis patients diagnosed within the study period times the length of the study period per 100 patients. Percentages were calculated as the number of patients who at any point suffered the events relative to ulcerative colitis diagnosis.

To identify differences between ulcerative colitis severity degrees, categorical variables were presented as frequencies and percentages. Continuous variables were summarized using means and standard deviations (SD) or medians and interquartile ranges (IQR), depending on the data distribution. The X^2^ or Fisher’s exact test was used to compare categorical variables. The Kruskal–Wallis test was used to compare continuous variables. All analyses were two-sided, and statistical significance was set at p < 0.05. Multiple comparison correction was carried with the Bonferroni method.

All analyses were carried out using Numpy (v2.1.1) [24] and SciPy (v1.14.1) [25] and statsmodels (v0.14.3) [26] through the TableOne package (v0.9.1) [27] using Python (v3.10.12). Visualizations were generated with seaborn (v0.13.2) [28].

### Secure data management

The data management circuit was designed to minimize the relative risk as described in the Impact Assessment on Data Protection analysis [29] and following the regulation for the use of medical data for research purposes in Andalusia (Joint Resolution 1/2021 of the General Secretariat for Research, Development and Innovation in Health of the Regional Ministry of Health and Families and the Management Directorate of the Andalusian Health Service [30]). Briefly, the data corresponding to the study approved by the ethics committee is requested to BPS. There, the data is extracted and pseudonymized by BPS personnel and then transferred to the iRWD [31, 32], a Secure Processing Environment (SPE) specifically designed for the secure analysis of data protected by the GDPR, being compliant with the definition of SPE as described in Article 50 of the Resolution of the European Parliament of 24 April 2024 on the proposal for a Regulation of the European Parliament and of the Council on the EHDS [33].

## Results

### Epidemiology of ulcerative colitis in Andalusia

A total of 23,518 adults with an ulcerative colitis diagnosis defined as described in Material and Methods, were found during the study period, of which 10,709 had their first diagnosis within the study period. Among these, a total of 4,810 patients with moderate to severe ulcerative colitis, defined by a series of treatment prescriptions (see Material and Methods) after the diagnosis of the disease, were found.

Table 1 describes the study population of patients with moderate to severe ulcerative colitis, which comprised a balanced distribution of sexes, with 54% men and 46% women, highlighting a relatively equal representation. The median age of patients was 43.7 years, with an interquartile range (IQR) from 32.1 to 57.4 years, indicating a middle-aged cohort. The median duration of ulcerative colitis was 5.9 years (IQR 3.5 to 8.1 years), suggesting a considerable duration of disease among the participants. The duration of disease was less than six years for 51.6% of patients, while 48.4% had been managing ulcerative colitis for six years or more, underscoring the chronic nature of the disease in nearly half of the population.

**Table 1:** Baseline demographic characteristics of patients with moderate to severe ulcerative colitis.

| Feature | Class | Value |
| --- | --- | --- |
| Sex (%) | Man | 2,595 (54.0%) |
|  | Woman | 2,215 (46.0%) |
| Age | Median [Q1, Q3] | 43.7 [32.1, 57.4] |
| Duration (years) | Median [Q1, Q3] | 5.9 [3.5, 8.1] |
| Disease duration (%) | <6 years | 2481 (51.6%) |
|  | >=6 years | 2329 (48.4%) |
| Treatment (%) | Corticoids | 3,123 (64.9%) |
|  | Immunosuppressor | 825 (17.2%) |
|  | Biological treatment | 862 (17.9%) |

Treatment regimens were dominated by corticosteroid use, reported in 64.9% of patients, reflecting their widespread use in managing ulcerative colitis flares. Immunosuppressive therapies were used by 17.2% of the cohort, while biological treatments were administered to 17.9%, indicating a significant reliance on advanced therapies for a subset of patients.

These findings provide a comprehensive overview of the demographic and clinical characteristics of the study population, emphasizing the diversity in disease severity and treatment approaches among patients with ulcerative colitis. The data also underscore the chronic nature of ulcerative colitis and the varied therapeutic strategies employed to manage this condition in a real-world setting. This analysis is crucial for understanding the risks of VTE, MACE, neoplasia, and serious infections in UC patients, informing future research and clinical practice.

### Incidence of associated diseases in ulcerative colitis patients

Ulcerative colitis is associated with an increased incidence of various comorbid conditions, including VTE, MACE, malignancies, and severe infections. The data from our study reveal that the incidence rates of these conditions vary significantly with age and time relative to the diagnosis of ulcerative colitis.

The incidence of VTE, encompassing both DVT and PE, is markedly higher in ulcerative colitis patients compared to the general population, that ranges from 0.10 to 0.18 per 100 individuals of European ancestry [34], especially among those aged over 60 years. This increased risk persists before and after the ulcerative colitis diagnosis, highlighting the chronic pro-inflammatory state and hypercoagulability in these patients. Similarly, MACE, including acute myocardial infarction and heart failure, show a notable rise with advancing age, reaching the highest incidence in patients older than 60 years.

Malignancies, excluding non-melanoma skin cancer (NMSC), are also more prevalent among ulcerative colitis patients, particularly those aged over 60 years. The risk of severe infections, such as bacterial pneumonia and sepsis, is also considerably elevated. These findings underscore the need for vigilant monitoring and proactive management of these comorbid conditions in ulcerative colitis patients to mitigate associated risks.

Table 2 outlines the incidence rates of associated diseases by age group, illustrating the heightened risk of thromboembolic events, cardiovascular conditions, malignancies, and severe infections in UC patients.

**Table 2:** Incidence of associated diseases and complications per 100 patients/year in moderate/severe ulcerative colitis patients by age group. Cells show incidences before / after / before and after the ulcerative colitis diagnosis

| Complication | 18-30 | 31-40 | 41-60 | >60 |
| --- | --- | --- | --- | --- |
| Deep vein thrombosis | 0.0 / 0.03 / 0.03 | 0.03 / 0.2 / 0.23 | 0.15 / 0.53 / 0.68 | 0.08 / 1.11 / 1.19 |
| Venous thromboembolism | 0.0 / 0.21 / 0.21 | 0.03 / 0.24 / 0.26 | 0.22 / 1.07 / 1.29 | 0.37 / 2.6 / 2.95 |
| Pulmonary embolism | 0.0 / 0.18 / 0.18 | 0.0 / 0.03 / 0.03 | 0.05 / 0.5 / 0.55 | 0.28 / 1.28 / 1.55 |
| MACE | 0.02 / 0.06 / 0.08 | 0.05 / 0.12 / 0.17 | 0.33 / 0.8 / 1.09 | 0.94 / 2.68 / 3.55 |
| Malignancies<br>(excluding NMSC) | 0.02 / 0.15 / 0.17 | 0.06 / 0.26 / 0.31 | 0.42 / 0.65 / 1.03 | 1.2 / 1.2 / 2.34 |
| Serious infection | 0.41 / 0.77 / 1.15 | 0.25 / 0.72 / 0.93 | 0.36 / 0.91 / 1.21 | 0.67 / 1.88 / 2.48 |
| Hospitalization | 0.25 / 0.47 / 0.7 | 0.17 / 0.44 / 0.59 | 0.27 / 0.69 / 0.93 | 0.52 / 0.96 / 1.44 |

A detailed analysis of the main hospital admission diagnoses due to infections among moderate/severe ulcerative colitis patients (Supplementary Figure 1) highlights that the proportion of hospitalizations due to infections increases significantly after the diagnosis of ulcerative colitis compared to the period before diagnosis. This trend suggests that ulcerative colitis, along with its associated treatments, such as corticoids, immunosuppressants and biologics, may elevate the susceptibility to severe infections. It also emphasizes the chronic and recurring nature of infectious complications in ulcerative colitis patients, which remain prevalent regardless of the timing of diagnosis. Interestingly, the predominant type of infection changes with the age of the patients, being predominant bacterial intestinal infections at younger ages, while pneumonia becomes prevalent for patients 40 years and older.

Supplementary Figure 2 illustrates the distribution of malignant neoplasms (non-CPNM) among patients with moderate/severe ulcerative colitis, broken down by whether the malignancy was diagnosed strictly before the UC diagnosis, strictly after, or at any time relative to the diagnosis (both before and after). The figure shows that a significant proportion of neoplasms are diagnosed after the UC diagnosis, indicating a potential link between ulcerative colitis and an increased risk of developing malignancies. This may be due to chronic inflammation, immune dysregulation, and the use of immunosuppressive therapies in ulcerative colitis management. However, neoplasms diagnosed before the onset of ulcerative colitis suggest that some patients may have pre-existing cancer risk factors independent of UC. Again, the dominant neoplasm type is clearly age-dependent.

The distribution of different types of DVT experienced by patients with moderate/severe ulcerative colitis, segmented by whether the thrombosis occurred strictly before the diagnosis, strictly after, or at any time relative to the diagnosis (both before and after) is shown in Supplementary Figure 3. The figure highlights that a substantial proportion of DVT cases occur after the diagnosis of ulcerative colitis, suggesting a strong association between the disease and an increased risk of thrombotic events. This increased risk is likely due to the chronic inflammation, altered coagulation pathways, and potential side effects of ulcerative colitis treatments, which are known to promote hypercoagulability. Like in neoplasms, the dominant DVT type changes with age, being more unspecific at younger ages and growing DVT associated to lower extremities at older ages.

Finally, the distribution of MACE events experienced by patients with moderate/severe ulcerative colitis with respect to the diagnosis (Supplementary Figure 4) indicates that the majority of MACE, including myocardial infarction, stroke, and heart failure, occur more frequently after the diagnosis of ulcerative colitis. This pattern suggests a possible link between ulcerative colitis and an increased risk of cardiovascular complications, which may be exacerbated by chronic inflammation, oxidative stress, and the adverse effects of long-term use of corticosteroids and other immunosuppressive medications commonly used in ulcerative colitis management. The higher incidence of MACE following ulcerative colitis diagnosis emphasizes the critical need for cardiovascular risk assessment and monitoring in these patients. Unlike in the previous cases, the prevalence of MACE type seems to be quite constant across the age range studied.

The analysis reveals that thromboembolic complications, particularly DVT and VTE, are significantly more prevalent in patients with moderate to severe ulcerative colitis compared to those with mild disease (see Table 3 and Supplementary Figure 1), while pulmonary embolism and MACE showed no significant difference in incidence across severity groups (see also Supplementary Figure 2). These findings highlight the increased thrombotic risk associated with disease severity, underscoring the importance of thromboprophylaxis and comprehensive risk assessment in managing UC patients, particularly those with moderate to severe disease.

**Table 3.**
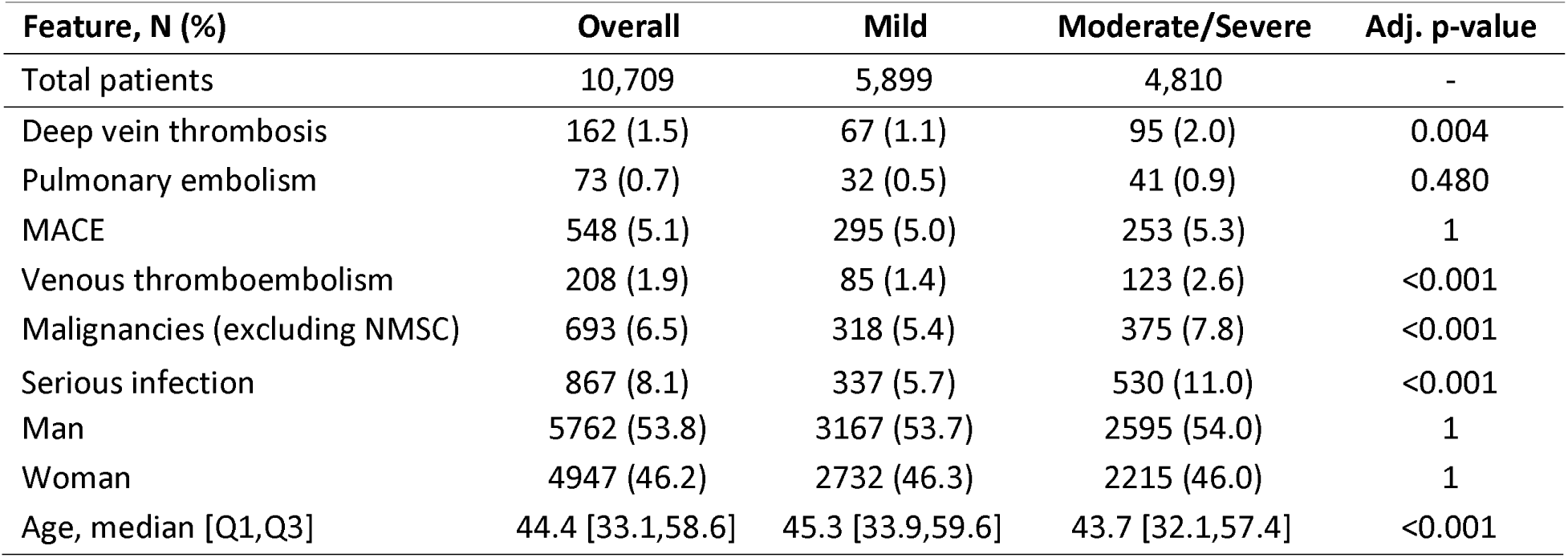
Occurrence of several features, including associated diseases and complications in mild and moderate to severe ulcerative colitis. The last column shows the X^2^ Bonferroni-adjusted p-value of the comparison of the distribution between both cases (except for the age, in which a Kruskal-Wallis test has been conducted and the non-adjusted p-value is shown).

| Feature, N (%) | Overall | Mild | Moderate/Severe | Adj. p-value |
| --- | --- | --- | --- | --- |
| Total patients | 10,709 | 5,899 | 4,810 | - |
| Deep vein thrombosis | 162 (1.5) | 67 (1.1) | 95 (2.0) | 0.004 |
| Pulmonary embolism | 73 (0.7) | 32 (0.5) | 41 (0.9) | 0.480 |
| MACE | 548 (5.1) | 295 (5.0) | 253 (5.3) | 1 |
| Venous thromboembolism | 208 (1.9) | 85 (1.4) | 123 (2.6) | <0.001 |
| Malignancies (excluding NMSC) | 693 (6.5) | 318 (5.4) | 375 (7.8) | <0.001 |
| Serious infection | 867 (8.1) | 337 (5.7) | 530 (11.0) | <0.001 |
| Man | 5762 (53.8) | 3167 (53.7) | 2595 (54.0) | 1 |
| Woman | 4947 (46.2) | 2732 (46.3) | 2215 (46.0) | 1 |
| Age, median [Q1,Q3] | 44.4 [33.1,58.6] | 45.3 [33.9,59.6] | 43.7 [32.1,57.4] | <0.001 |

Also, the incidence of malignancies (excluding NMSC), including breast, lung, colon prostate and other (see Supplementary Figure 3) and serious infections (see Supplementary Figure 4) is significantly higher in moderate to severe than in mild ulcerative colitis.

Man and woman show no significantly different distributions across ulcerative colitis severity, while the median age of diagnosis was significantly higher in mild ulcerative colitis than in moderate to severe disease.

### Risk factors in ulcerative colitis

Table 4 presents the percentages of the total number of moderate/severe ulcerative colitis patients with various cardiovascular and metabolic risk factors, categorized both by their occurrence relative to the ulcerative colitis diagnosis and by age range. Note that patients can have more than one risk factor or extraintestinal manifestation and diverse occurrences at different relative times with respect to the diagnosis.

**Table 4.** Percentages of the total number of moderate/severe ulcerative colitis patients with different cardiovascular and metabolic risk factors, categorized by age range. Each cell represents percentages of occurrence relative to the ulcerative colitis diagnosis: before/after/before and after.

| Risk factor | 18-30 | 31-40 | 41-60 | >60 |
| --- | --- | --- | --- | --- |
| Obesity | 1.6 / 1.3 / 2.9 | 1.7 / 2.8 / 4.4 | 3.5 / 3.9 / 6.9 | 5.5 / 4.7 / 10.1 |
| Hypertension | 1.0 / 1.5 / 2.4 | 3.1 / 5.0 / 7.9 | 17.9 / 12.3 / 29.3 | 46.3 / 13.7 / 58.1 |
| Hypercholesterolemia | 0.4 / 0.9 / 1.3 | 1.5 / 3.2 / 4.7 | 6.1 / 7.9 / 13.7 | 9.9 / 7.5 / 16.8 |
| Hormonal treatment | 1.5 / 6.9 / 6.9 | 1.0 / 6.8 / 6.8 | 0.4 / 4.5 / 4.5 | 0.9 / 4.1 / 4.1 |
| Coronary artery disease | 0.0 / 0.2 / 0.5 | 0.3 / 0.2 / 0.4 | 0.8 / 1.8 / 3.5 | 2.4 / 4.5 / 9.8 |
| Tobacco dependence | 2.5 / 6.0 / 7.4 | 4.6 / 7.0 / 10.1 | 7.4 / 6.5 / 12.8 | 6.1 / 5.5 / 11.0 |
| Diabetes | 1.2 / 1.1 / 2.1 | 1.4 / 1.8 / 3.2 | 7.2 / 6.5 / 13.1 | 20.6 / 8.1 / 27.8 |
| Heart failure | 0.1 / 0.0 / 0.1 | 0.2 / 0.0 / 0.2 | 0.8 / 1.7 / 2.4 | 6.7 / 11.6 / 17.2 |
| Ischemic heart disease | 0.0 / 0.1 / 0.1 | 0.2 / 0.0 / 0.2 | 1.8 / 1.4 / 3.1 | 7.3 / 4.7 / 11.7 |
| Thrombophilia | 0.0 / 0.3 / 0.3 | 0.2 / 0.9 / 1.1 | 0.1 / 0.4 / 0.5 | 0.2 / 0.8 / 1.0 |
| Atrioventricular block | 0.0 / 0.0 / 0.0 | 0.0 / 0.1 / 0.1 | 0.0 / 0.2 / 0.2 | 0.7 / 1.9 / 2.4 |
| Cardiac arrest | 0.0 / 0.0 / 0.0 | 0.1 / 0.0 / 0.1 | 0.1 / 0.1 / 0.1 | 0.1 / 0.1 / 0.2 |

The data reveal that risk factors are systematically higher in older age groups (>60 years) and tend to increase after the diagnosis of ulcerative colitis, indicating the compounding effect of chronic inflammation and ulcerative colitis treatments on cardiovascular health. The most significant risk factor identified is hypertension, particularly among patients aged over 60, although hypercholesterolemia and heart failure, with 16.8% and 17.2% of patients over 60 affected, respectively, are also remarkable. There is also a high prevalence of diabetes and coronary artery disease among older patients.

Contrarily, obesity and tobacco dependency are prevalent across all age groups, underscoring the need for lifestyle interventions.

As expected, age is a critical determinant of risk factor prevalence. Older patients (over 60 years) exhibit a higher incidence of comorbidities such as hypertension, hypercholesterolemia, and diabetes compared to younger age groups. This trend emphasizes the cumulative effect of aging and chronic ulcerative colitis on the cardiovascular system. Younger patients, while still at risk, generally have fewer comorbid conditions, which correlates with a slightly lower cardiovascular risk profile.

### Autoimmune comorbidities in ulcerative colitis

Table 5 presents the percentages of the total number of moderate/severe ulcerative colitis patients with several autoimmune comorbidities, categorized both by their occurrence relative to the ulcerative colitis diagnosis and by age range. It also includes the reported prevalence of each autoimmune condition in the general population. Note that patients can have more than one occurrence at different relative times with respect to the diagnosis.

**Table 5.**
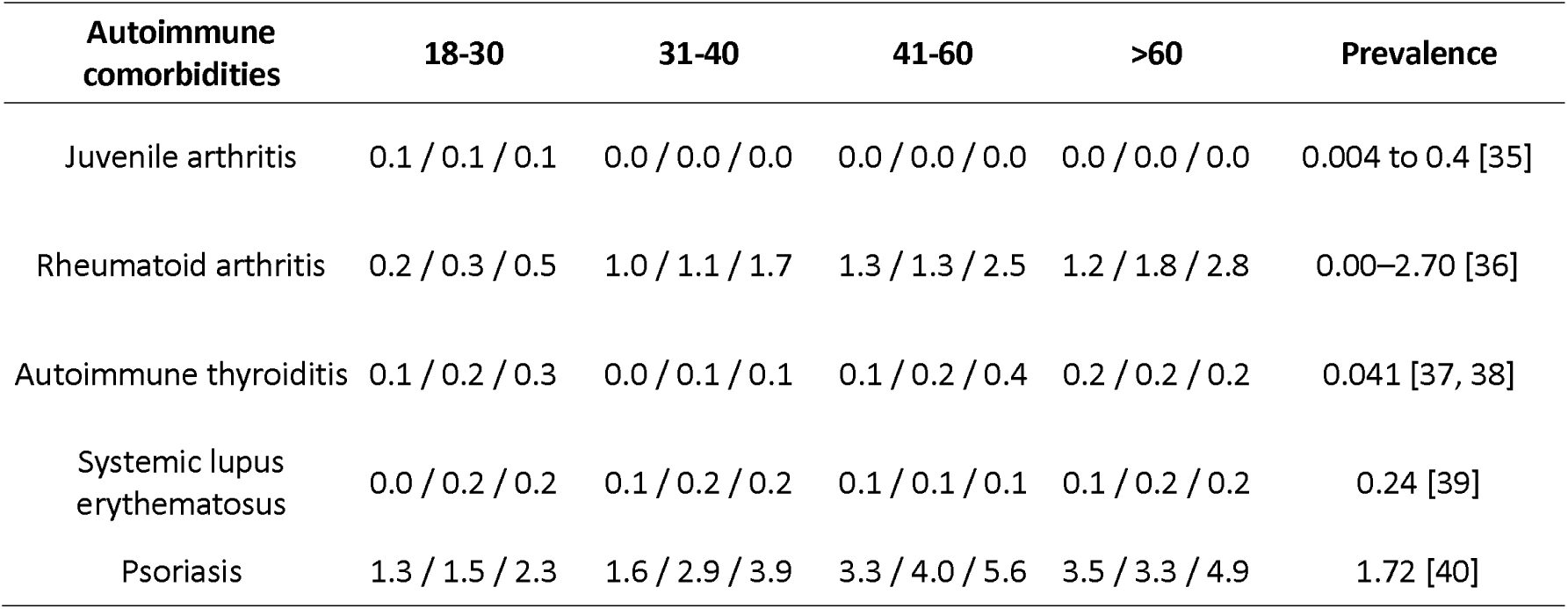
Percentages of the total number of moderate/severe ulcerative colitis patients with different autoimmune comorbidities, categorized by age range. Each column shows percentages of occurrence relative to the ulcerative colitis diagnosis: before/after/before and after.

| Autoimmune comorbidities | 18-30 | 31-40 | 41-60 | >60 | Prevalence |
| --- | --- | --- | --- | --- | --- |
| Juvenile arthritis | 0.1 / 0.1 / 0.1 | 0.0 / 0.0 / 0.0 | 0.0 / 0.0 / 0.0 | 0.0 / 0.0 / 0.0 | 0.004 to 0.4 [35] |
| Rheumatoid arthritis | 0.2 / 0.3 / 0.5 | 1.0 / 1.1 / 1.7 | 1.3 / 1.3 / 2.5 | 1.2 / 1.8 / 2.8 | 0.00–2.70 [36] |
| Autoimmune thyroiditis | 0.1 / 0.2 / 0.3 | 0.0 / 0.1 / 0.1 | 0.1 / 0.2 / 0.4 | 0.2 / 0.2 / 0.2 | 0.041 [37, 38] |
| Systemic lupus erythematosus | 0.0 / 0.2 / 0.2 | 0.1 / 0.2 / 0.2 | 0.1 / 0.1 / 0.1 | 0.1 / 0.2 / 0.2 | 0.24 [39] |
| Psoriasis | 1.3 / 1.5 / 2.3 | 1.6 / 2.9 / 3.9 | 3.3 / 4.0 / 5.6 | 3.5 / 3.3 / 4.9 | 1.72 [40] |

The percentage of patients with autoimmune comorbidities is low comparatively to the case of risk factors. There is also some influence of the age group in the prevalence of the conditions (in the case of juvenile arthritis the effect is the opposite). For Juvenile arthritis, rheumatoid arthritis and systemic lupus erythematosus, the observed percentages are similar to the respective prevalence described for general population, represented in the last column of

Table 5. However, the incidence of Autoimmune thyroiditis seems to be higher in ulcerative patients than in general population and, similarly in the case of psoriasis at older ages.

### Hormonal treatments in ulcerative colitis patients

The type of hormonal treatment in moderate/severe ulcerative colitis patients is markedly age-dependent (see Figure 1), with contraceptives being the most common in reproductive-age individuals. This shifts to hormone replacement therapy becoming more prevalent in patients over 40, and by age 60, it is typically the only hormonal treatment used. Overall, the total number of hormonal treatments decreases as patients age.

**Figure 1.**
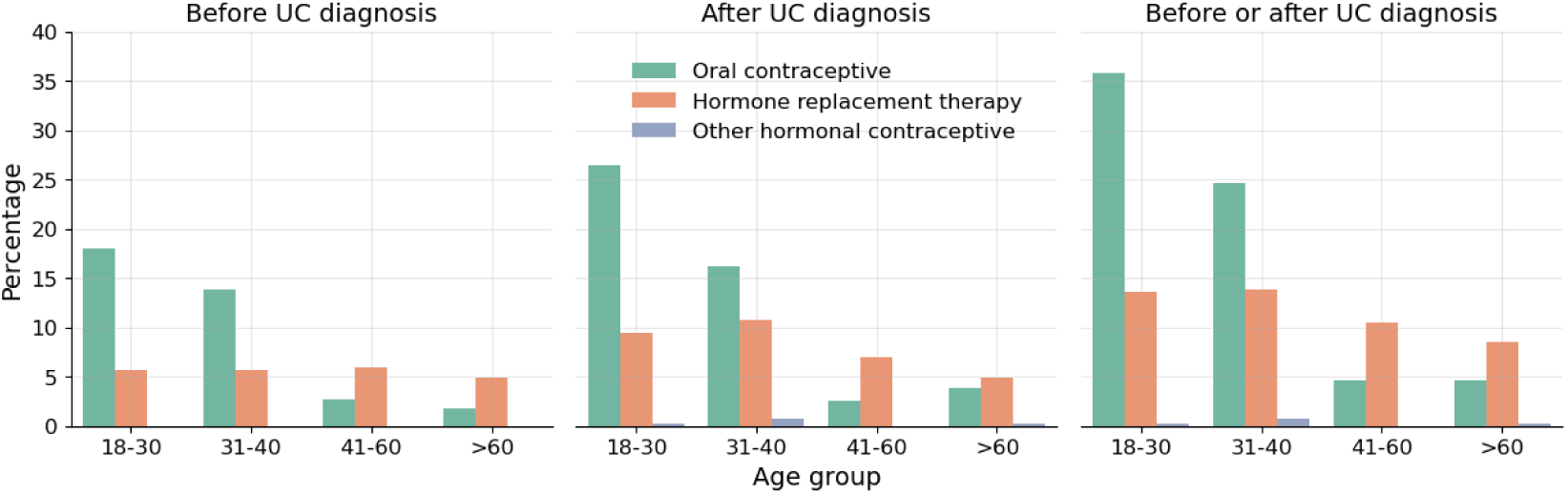
Percentages of types of hormonal treatments prescribed to ulcerative colitis patients, stratified according to whether the date of dispensing the drug occurred strictly before or after the diagnosis, or either before or after.

Irrespective of the type of hormonal treatment, it is always higher after the diagnosis of ulcerative colitis than before.

### Hospitalization in ulcerative colitis patients

Table 6 provides a detailed analysis of hospitalization incidences before, after, and both before and after the ulcerative colitis diagnosis in patients of different age groups.

**Table 6:** Incidence per 100 patients/year of hospitalizations patients of different age group with ulcerative colitis, stratified according to whether the date of hospitalization occurred strictly before or after the diagnosis, or either before or after.

| Age group | Before UC diagnosis | After UC diagnosis | Before and after UC diagnosis |
| --- | --- | --- | --- |
| 18-30 | 0.25 | 0.47 | 0.7 |
| 31-40 | 0.17 | 0.44 | 0.59 |
| 41-60 | 0.27 | 0.69 | 0.93 |
| >60 | 0.52 | 0.96 | 1.44 |

The data show a clear increase in hospitalization rates after the diagnosis of ulcerative colitis. The post-diagnosis incidence is remarkably higher, likely reflecting the complications associated with managing a chronic inflammatory disease.

## Discussion

This study aimed to provide a comprehensive analysis of the risks associated with VTE, MACE, malignancies, and serious infections in patients with ulcerative colitis using real-world data from a total of 23,518 patients from the Andalusian BPS. The findings highlight several remarkable trends and underline the importance of proactive management in ulcerative colitis patients to mitigate these risks.

The elevated incidence of VTE and MACE among ulcerative colitis patients with respect to observations reported for the general population [34], particularly those over 60 years of age, is a major concern [41]. Chronic inflammation, which characterizes this disease, contributes to hypercoagulability, leading to an increased risk of thrombotic events. The incidence of VTE, including both DVT and pulmonary embolism, was notably higher in ulcerative colitis patients compared to the general population, with rates increasing significantly after the diagnosis of the disease [42]. This trend underscores the need for early cardiovascular risk assessment and the implementation of thromboprophylaxis strategies, particularly during hospitalization or active disease phases.

The association between ulcerative colitis and MACE was similarly concerning. Our findings revealed a marked increase in the incidence of MACE, such as myocardial infarction and heart failure, especially in older patients. Chronic inflammation and the long-term use of corticosteroids and other immunosuppressive therapies likely contribute to this elevated risk. Given the significant cardiovascular burden identified in this study, routine cardiovascular monitoring and the management of modifiable risk factors, such as hypertension and hyperlipidemia, are crucial in the care of UC patients.

The study also revealed a higher prevalence of malignancies, particularly in patients over 60 years of age. The link between ulcerative colitis and malignancy, especially colorectal cancer, has been well-documented [43], and our findings reinforce the need for vigilant cancer surveillance in this population. Immunosuppressive treatments, commonly used to manage ulcerative colitis, may contribute to an increased cancer risk in inflammatory diseases, particularly with long-term use [44]. Therefore, early detection and intervention remain critical in reducing cancer-related morbidity and mortality in UC patients.

Infection-related hospitalizations were another significant finding of this study. ulcerative colitis patients, especially those receiving immunosuppressive treatments, exhibited a higher incidence of severe infections, including bacterial pneumonia and sepsis. The chronic nature of ulcerative colitis and the frequent use of biologics and corticosteroids increase the susceptibility to infections [45]. This highlights the importance of preventive measures, such as vaccination and routine infection screening, in reducing infection-related complications in UC patients.

Interestingly, a smaller but notable number of hospitalizations occur before the UC diagnosis, which may be linked to misdiagnosed gastrointestinal symptoms or early manifestations of the disease that were not identified as ulcerative colitis initially (see Table 4). Hospitalizations before and after diagnosis suggests ongoing medical needs in patients who may have experienced health issues related to ulcerative colitis before their formal diagnosis and continue to require medical intervention after. This emphasizes the importance of close medical management both pre-and post-diagnosis to mitigate complications and hospitalizations

Another important aspect derived from this study is related the potential impact of the observations made in prevention. The results presented suggest that ulcerative colitis affects individuals during their most productive years, underscoring the impact of the disease on personal and professional aspects of life. Additionally, the relatively broad age range at diagnosis emphasizes the need for vigilance across a wide age spectrum in clinical settings, as the disease can present variably depending on age at onset. This distribution pattern aligns with existing literature, which indicates that ulcerative colitis frequently occurs in young adults but remains a considerable concern for older populations as well. It also underscores the need for careful monitoring and preventive measures to reduce infection-related hospitalizations in ulcerative colitis patients, particularly in those receiving immunosuppressive therapies. Understanding the temporal patterns of infection-related hospitalizations can inform clinical decisions and guide strategies to minimize these complications in ulcerative colitis management.

The observed distribution of malignant neoplasms after the ulcerative colitis diagnosis (Supplementary Figure 2) emphasizes the importance of cancer surveillance in patients, especially for those receiving long-term immunosuppressive or biologic treatments. Early detection and proactive management of cancer risks are crucial in this population to improve outcomes and reduce the burden of malignancies associated with ulcerative colitis. With respect to DTV, data from Supplementary Figure 3 underscore the need for preventive measures, including thromboprophylaxis, especially during active disease phases or hospitalization periods. Understanding the timing and types of DVT in relation to ulcerative colitis diagnosis can guide clinicians in tailoring risk-reducing strategies to minimize thrombotic complications in this vulnerable patient group.

Table 3 shows a high prevalence of diabetes and coronary artery disease among older patients, which emphasizes the necessity for comprehensive cardiovascular risk management in ulcerative colitis patients, particularly those with prolonged disease duration and exposure to corticosteroids or immunosuppressive therapies. The findings suggest that continuous monitoring and early intervention for these risk factors are crucial to reduce the burden of cardiovascular diseases in such patients. Obesity and smoking dependency are also prevalent across all age groups, underscoring the need for lifestyle interventions.

Another interesting observation from table 3 is the significantly higher incidence of VTE, malignancies and serious infections as the severity of ulcerative colitis increases. This is probably due to the use of immunosuppressive therapies, which are associated with an increased risk of infections, and likely with VTE and malignancies.

It is worth noting that some autoimmune conditions, such as autoimmune thyroiditis, or psoriasis at old ages, seem to occur more frequently in ulcerative colitis patients than in general population (see Table 4). Although a higher incidence of psoriasis in ulcerative colitis patients has already been described [46], a higher incidence of autoimmune thyroiditis in ulcerative colitis patients had not been reported to date, although it was suggested by studies that describe higher prevalence of hypothyroidism in ulcerative colitis patients with respect to the normal population [47], while others suggest that Crohn’s disease, but not ulcerative colitis, is a risk factor for autoimmune thyroiditis [48].

The use of RWD from the Andalusian BPS was instrumental in providing a broad and detailed analysis of the risks associated with ulcerative colitis. RWD offers a unique opportunity to capture a more comprehensive picture of patient outcomes by utilizing data from routine clinical settings, rather than controlled trial environments [49]. Andalusia, with its large population and extensive BPS resource, stands at the forefront of this data revolution. The BPS not only centralizes patient data across the region but also facilitates large-scale, population-based studies that can inform health policies and improve clinical practices [19]. As healthcare systems generate more data, the potential for future studies expands significantly. This wealth of information will enable researchers to identify trends, refine risk models, and explore new avenues for personalized medicine and targeted therapies, particularly for chronic conditions like ulcerative colitis.

However, while the study capitalized on these strengths, several limitations remain. The retrospective design may have introduced selection bias, and reliance on electronic health records can sometimes result in underreporting or misclassification of certain conditions. Furthermore, lifestyle factors such as diet and physical activity were not explored in this analysis. As more medical data becomes available, future studies can address these gaps, providing a more holistic understanding of disease management and outcomes.

In conclusion, the findings from this study highlight the significant cardiovascular, thromboembolic, oncological, and infectious risks associated with ulcerative colitis, particularly in older patients. Proactive management, including regular cardiovascular monitoring, cancer surveillance, and infection prevention strategies, is essential in improving outcomes for ulcerative colitis patients. Future research should focus on exploring the impact of lifestyle interventions and developing tailored strategies to mitigate these risks in ulcerative colitis patients. This study underscores the value of real-world data in guiding clinical decision-making and improving patient care in ulcerative colitis.

## Ethics statement

The Andalusian Biomedical Research Ethics Coordinating Committee approved the study entitled: “Analysis of real-world data on venous thromboembolism, major adverse cardiovascular events, neoplasms and serious infections in the Andalusian Population Health Database to improve the management of patients with ulcerative colitis and atopic dermatitis”; (March 28, 2023, Acta 03/23) and waived informed consent for the secondary use of clinical data for research purposes.

## Data sharing

The datasets analyzed during the current study are available in the Population Health Database repository (https://www.sspa.juntadeandalucia.es/servicioandaluzdesalud/profesionales/sistemas-de-informacion/base-poblacional-de-salud), subject to controlled access according to the regulation for the use of medical data for research in the Andalusian Health System (https://www.sspa.juntadeandalucia.es/servicioandaluzdesalud/sites/default/files/sincfiles/wsas-media-sas_normativa_mediafile/2021/resolucion_conjunta_acceso_a_datos_investigacion_def_20211201%28F%29.pdf).

## Acknowledgements

This work has been funded by Pfizer quality improvement project entitled “Real world data analysis of venous thromboembolism, major adverse cardiovascular events, neoplasia and severe infections in the Andalusian Population Health Database for the improvement of the management of patients with ulcerous colitis and atopic dermatitis” (77644553) and co-financed by the Consejeria de Salud y Consumo, Junta de Andalucia (IE19_259 FPS).

