## Supplementary Table 1 for "Real world data analysis of venous thromboembolism, adverse major cardiovascular events, neoplasia and serious infections in ulcerative colitis patients"

### SUPPLEMENTARY MATERIAL

**Supplementary Table 1:** ICD-10 codes used in the study

| Adverse events | ICD-10 | Description |
|---|---|---|
| Deep vein thrombosis (DVT) | I26.09 | Other pulmonary embolism with acute cor pulmonale |
| | I26.90 | Septic pulmonary embolism without acute cor pulmonale |
| | I26.99 | Other pulmonary embolism without acute cor pulmonale |
| Pulmonary embolism (PE) | I82.0 | Budd-Chiari syndrome |
| | I82.1 | Thrombophlebitis migrans |
| | I82.2 | Embolism and thrombosis of vena cava and other thoracic veins |
| | I82.3 | Embolism and thrombosis of renal vein |

| | | |
|---|---|---|
| | I82.4 | Acute embolism and thrombosis of deep veins of lower extremity |
| | I82.5 | Chronic embolism and thrombosis of deep veins of lower extremity |
| | I82.60 | Acute embolism and thrombosis of unspecified veins of upper extremity |
| | I82.62 | Acute embolism and thrombosis of deep veins of upper extremity |
| | I82.72 | Chronic embolism and thrombosis of deep veins of upper extremity |
| | I82.89 | Embolism and thrombosis of other specified veins |
| | I82.9 | Embolism and thrombosis of unspecified vein |
| | I82.A | Embolism and thrombosis of axillary vein |
| | I82.B | Embolism and thrombosis of subclavian vein |
| | I82.C | Embolism and thrombosis of internal jugular vein |
| Major cardiovascular events (MACE) | G45 | Transient cerebral ischemic attacks and related syndromes |
| | I21 | Acute myocardial infarction |
| | I46 | Cardiac arrest |
| | I50 | Heart failure |
| | I63 | Cerebral infarction |
| Malignant neoplasm except non-melanoma skin cancer | C07 | Malignant neoplasm of parotid gland |

| | |
|---|---|
| C15 | Malignant neoplasm of esophagus |
| C16 | Malignant neoplasm of stomach |
| C17 | Malignant neoplasm of small intestine |
| C18 | Malignant neoplasm of colon |
| C19 | Malignant neoplasm of rectosigmoid junction |
| C20 | Malignant neoplasm of rectum |
| C22 | Malignant neoplasm of liver and intrahepatic bile ducts |
| C24 | Malignant neoplasm of other and unspecified parts of biliary tract |
| C25 | Malignant neoplasm of pancreas |
| C26 | Malignant neoplasm of other and ill-defined digestive organs |
| C32 | Malignant neoplasm of larynx |
| C33 | Malignant neoplasm of trachea |
| C34 | Malignant neoplasm of bronchus and lung |
| C37 | Malignant neoplasm of thymus |
| C38 | Malignant neoplasm of heart, mediastinum and pleura |
| C43 | Malignant melanoma of skin |
| C44 | Other and unspecified malignant neoplasm of skin |

| | |
|---|---|
| C45 | Mesothelioma |
| C48 | Malignant neoplasm of retroperitoneum and peritoneum |
| C49 | Malignant neoplasm of other connective and soft tissue |
| C50 | Malignant neoplasm of breast |
| C51 | Malignant neoplasm of vulva |
| C60 | Malignant neoplasm of penis |
| C61 | Malignant neoplasm of prostate |
| C63 | Malignant neoplasm of other and unspecified male genital organs |
| C64 | Malignant neoplasm of kidney, except renal pelvis |
| C66 | Malignant neoplasm of ureter |
| C67 | Malignant neoplasm of bladder |
| C71 | Malignant neoplasm of brain |
| C75 | Malignant neoplasm of other endocrine glands and related structures |
| C76 | Malignant neoplasm of other and ill-defined sites |
| C77 | Secondary and unspecified malignant neoplasm of lymph nodes |
| C78 | Secondary malignant neoplasm of respiratory and digestive organs |
| C79 | Secondary malignant neoplasm of other and unspecified sites |

| | | |
|---|---|---|
| | C80 | Malignant neoplasm without specification of site |
| | C81 | Hodgkin lymphoma |
| | C83 | Non-follicular lymphoma |
| | C84 | Mature T/NK-cell lymphomas |
| | C85 | Other specified and unspecified types of non-Hodgkin lymphoma |
| | C90 | Multiple myeloma and malignant plasma cell neoplasms |
| | C91 | Lymphoid leukemia |
| | C92 | Myeloid leukemia |
| | C95 | Leukemia of unspecified cell type |
| | C96 | Other and unspecified malignant neoplasms of lymphoid, hematopoietic and related tissue |
| Infection | A02 | Other salmonella infections |
| | A04 | Other bacterial intestinal infections |
| | A05 | Other bacterial foodborne intoxications, not elsewhere classified |
| | A06 | Amebiasis |
| | A07 | Other protozoal intestinal diseases |
| | A08 | Viral and other specified intestinal infections |

| | |
|---|---|
| A09 | Infectious gastroenteritis and colitis, unspecified |
| A15 | Respiratory tuberculosis |
| A18 | Tuberculosis of other organs |
| A19 | Miliary tuberculosis |
| A27 | Leptospirosis |
| A31 | Infection due to other mycobacteria |
| A32 | Listeriosis |
| A37 | Whooping cough |
| A39 | Meningococcal infection |
| A40 | Streptococcal sepsis |
| A41 | Other sepsis |
| A43 | Nocardiosis |
| A46 | Erysipelas |
| A48 | Other bacterial diseases, not elsewhere classified |
| A49 | Bacterial infection of unspecified site |
| A50 | Congenital syphilis |
| A51 | Early syphilis |

| | |
|---|---|
| A52 | Late syphilis |
| A53 | Other and unspecified syphilis |
| A54 | Gonococcal infection |
| A55 | Chlamydial lymphogranuloma (venereum) |
| A56 | Other sexually transmitted chlamydial diseases |
| A59 | Trichomoniasis |
| A60 | Anogenital herpesviral [herpes simplex] infections |
| A63 | Other predominantly sexually transmitted diseases, not elsewhere classified |
| A70 | Chlamydia psittaci infections |
| A78 | Q fever |
| A79 | Other rickettsioses |
| A81 | Atypical virus infections of central nervous system |
| A85 | Other viral encephalitis, not elsewhere classified |
| A86 | Unspecified viral encephalitis |
| A87 | Viral meningitis |
| B00 | Herpesviral [herpes simplex] infections |
| B01 | Varicella [chickenpox] |

| | |
|---|---|
| B02 | Zoster [herpes zoster] |
| B08 | Other viral infections characterized by skin and mucous membrane lesions, not elsewhere classified |
| B09 | Unspecified viral infection characterized by skin and mucous membrane lesions |
| B15 | Acute hepatitis A |
| B16 | Acute hepatitis B |
| B17 | Other acute viral hepatitis |
| B18 | Chronic viral hepatitis |
| B19 | Unspecified viral hepatitis |
| B20 | Human immunodeficiency virus [HIV] disease |
| B25 | Cytomegaloviral disease |
| B26 | Mumps |
| B27 | Infectious mononucleosis |
| B34 | Viral infection of unspecified site |
| B35 | Dermatophytosis |
| B36 | Other superficial mycoses |
| B37 | Candidiasis |
| B44 | Aspergillosis |

| | |
|---|---|
| B45 | Cryptococcosis |
| B46 | Zygomycosis |
| B48 | Other mycoses, not elsewhere classified |
| B49 | Unspecified mycosis |
| B50 | Plasmodium falciparum malaria |
| B55 | Leishmaniasis |
| B57 | Chagas' disease |
| B59 | Pneumocystosis |
| B75 | Trichinellosis |
| B78 | Strongyloidiasis |
| B82 | Unspecified intestinal parasitism |
| B85 | Pediculosis and phthiriasis |
| B86 | Scabies |
| B87 | Myiasis |
| B89 | Unspecified parasitic disease |
| B90 | Sequelae of tuberculosis |
| B91 | Sequelae of poliomyelitis |

| | |
|---|---|
| B94 | Sequelae of other and unspecified infectious and parasitic diseases |
| B95 | Streptococcus, Staphylococcus, and Enterococcus as the cause of diseases classified elsewhere |
| B96 | Other bacterial agents as the cause of diseases classified elsewhere |
| B97 | Viral agents as the cause of diseases classified elsewhere |
| B99 | Other and unspecified infectious diseases |
| G00 | Bacterial meningitis, not elsewhere classified |
| I33 | Acute and subacute endocarditis |
| I96 | Gangrene, not elsewhere classified |
| J03 | Acute tonsillitis |
| J09 | Influenza due to certain identified influenza viruses |
| J10 | Influenza due to other identified influenza virus |
| J11 | Influenza due to unidentified influenza virus |
| J12 | Viral pneumonia, not elsewhere classified |
| J13 | Pneumonia due to Streptococcus pneumoniae |
| J14 | Pneumonia due to Hemophilus influenzae |
| J15 | Bacterial pneumonia, not elsewhere classified |
| J18 | Pneumonia, unspecified organism |

| | |
|---|---|
| J20 | Acute bronchitis |
| J35 | Chronic diseases of tonsils and adenoids |
| J47 | Bronchiectasis |
| J85 | Abscess of lung and mediastinum |
| K85 | Acute pancreatitis |
| K94 | Complications of artificial openings of the digestive system |
| M00 | Pyogenic arthritis |
| M46 | Other inflammatory spondylopathies |
| M60 | Myositis |
| N30 | Cystitis |
| O23 | Infections of genitourinary tract in pregnancy |
| O85 | Puerperal sepsis |
| R65 | Symptoms and signs specifically associated with systemic inflammation and infection |
| T80 | Complications following infusion, transfusion and therapeutic injection |
| T82 | Complications of cardiac and vascular prosthetic devices, implants and grafts |
| T83 | Complications of genitourinary prosthetic devices, implants and grafts |
| T84 | Complications of internal orthopedic prosthetic devices, implants and grafts |

| | |
|---|---|
| T85 | Complications of other internal prosthetic devices, implants and grafts |
| T86 | Complications of transplanted organs and tissue |
| T87 | Complications peculiar to reattachment and amputation |
