## Supplementary Figure 1 for "Real world data analysis of venous thromboembolism, adverse major cardiovascular events, neoplasia and serious infections in ulcerative colitis patients"

**SUPPLEMENTARY MATERIAL**

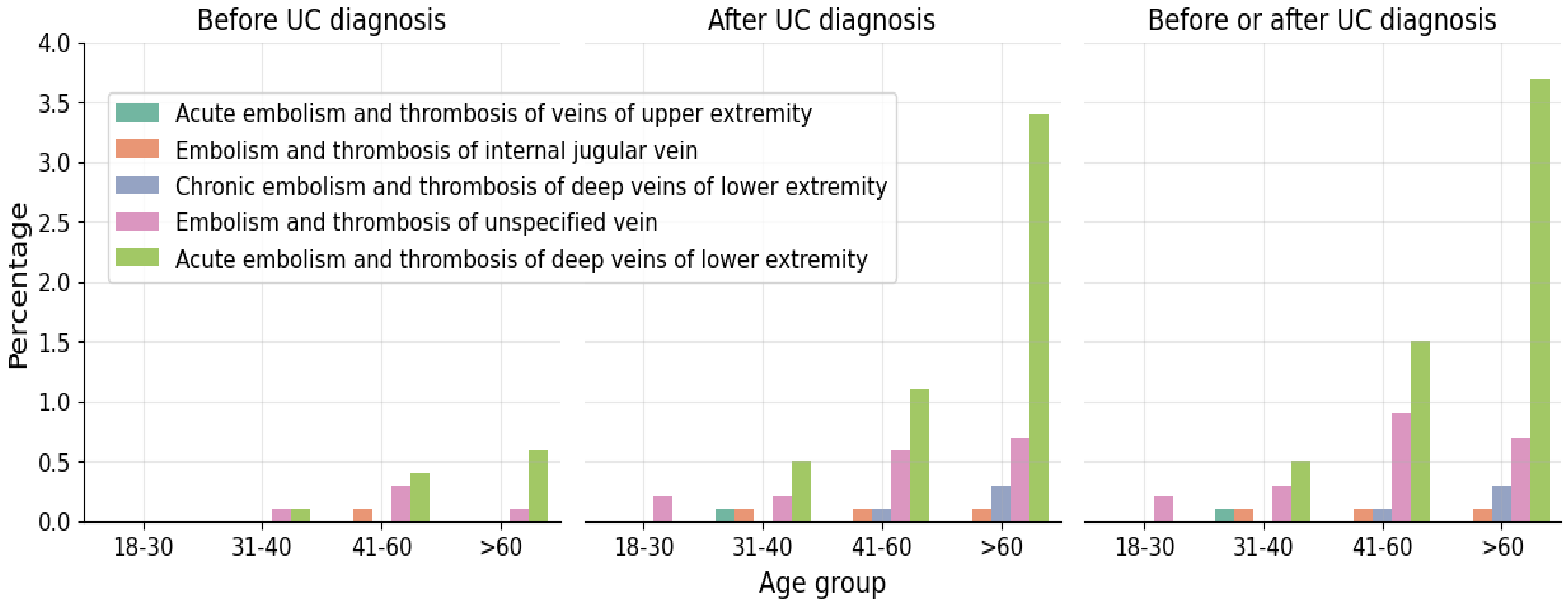


**Supplementary Figure 1:** Top 5 most prevalent DVT in patients according to age group and relative to ulcerative colitis diagnosis
