## Supplementary Figure 2 for "Real world data analysis of venous thromboembolism, adverse major cardiovascular events, neoplasia and serious infections in ulcerative colitis patients"

### SUPPLEMENTARY MATERIAL

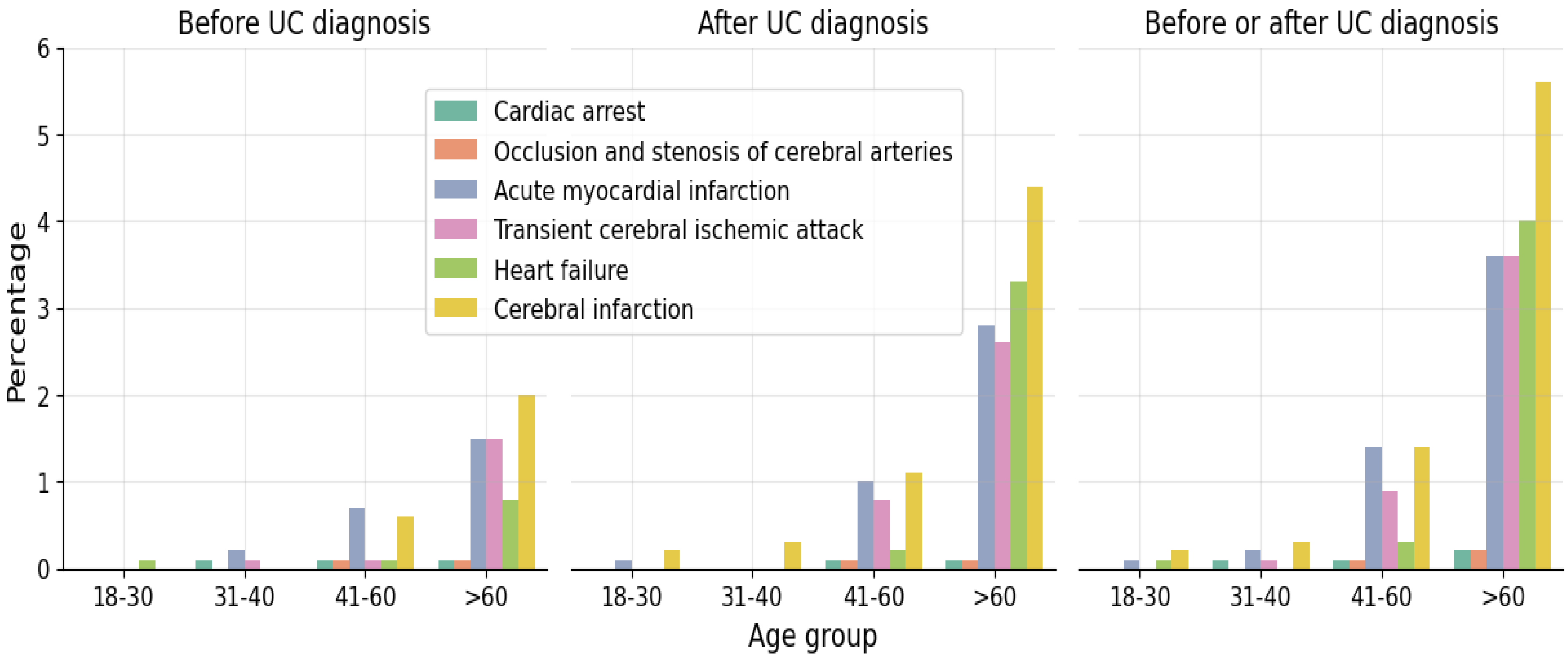


**Supplementary Figure 2:** Percentage of patients having with a MACE according to age group and relative to ulcerative colitis diagnosis
