## Supplementary Figure 4 for "Real world data analysis of venous thromboembolism, adverse major cardiovascular events, neoplasia and serious infections in ulcerative colitis patients"

### SUPPLEMENTARY MATERIAL

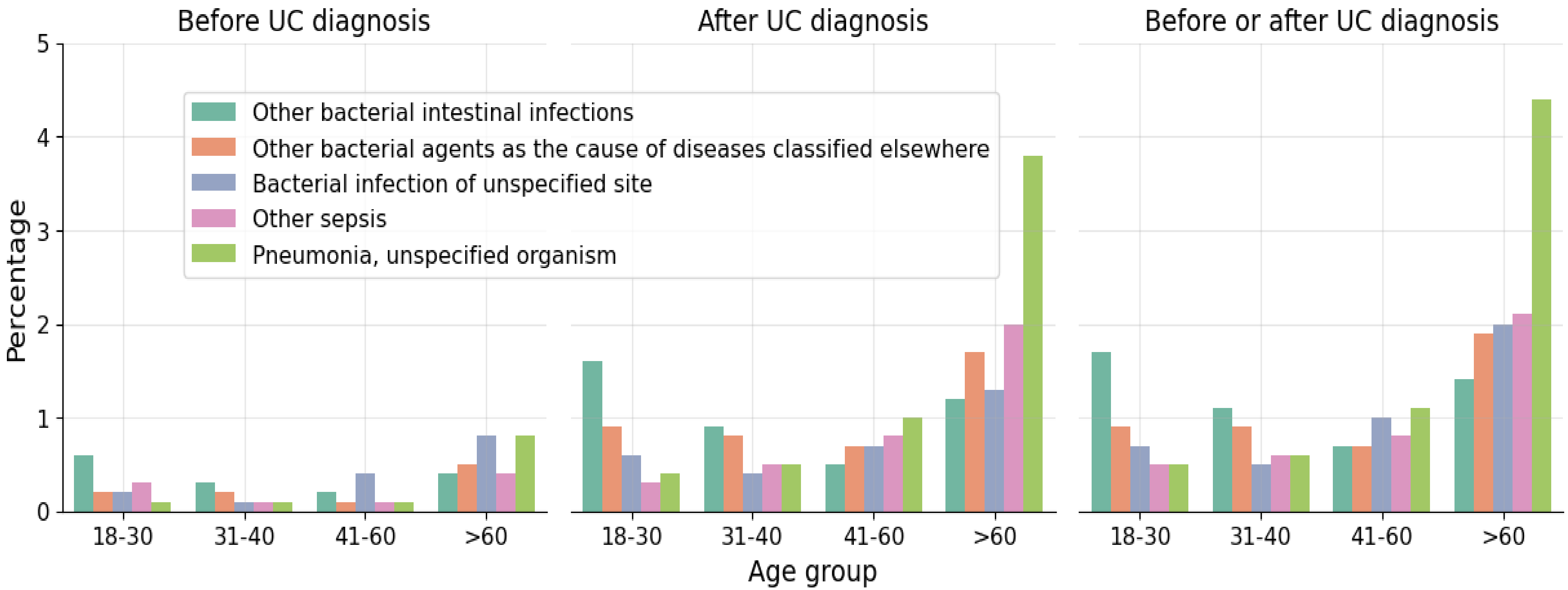


**Supplementary Figure 4:** Top 5 most prevalent bacterial infections leading to patient hospitalization according to age group and relative to ulcerative colitis diagnosis
